# Hyponatraemia and mortality in psychiatric patients: protocol for Bayesian causal inference study

**DOI:** 10.1101/2020.06.25.20138206

**Authors:** Benjamin Skov Kaas-Hansen, Niels Graudal, Stig Ejdrup Andersen, Lau Caspar Thygesen, Søren Brunak, Gesche Jürgens

**Affiliations:** Clinical Pharmacology Unit, Zealand University Hospital, Denmark; NNF Center for Protein Research, University of Copenhagen, Denmark; The Lupus and Vasculitis Clinic VRR 4242, Copenhagen University Hospital, Denmark; National Institute of Public Health, University of Southern Denmark, Denmark

**Keywords:** Hyponatraemia (https://meshb.nlm.nih.gov/record/ui?ui=D007010), Causality (https://meshb.nlm.nih.gov/record/ui?ui=D015984), Mortality (https://meshb.nlm.nih.gov/record/ui?ui=D009026), Bayesian Analysis (https://meshb.nlm.nih.gov/record/ui?ui=D001499), Antipsychotics (https://meshb.nlm.nih.gov/record/ui?ui=D014150), Psychopharmaceuticals (https://meshb.nlm.nih.gov/record/ui?ui=D011619), Polypharmacy (https://meshb.nlm.nih.gov/record/ui?ui=D019338)

## Abstract

**Introduction:** Hyponatriaemia has been associated with mortality in somatic patients, but little is known about the nature of such an association in psychiatric patients. This manuscript details a study to investigate if hyponatraemia be a direct contributor to death in schizophrenic and bipolar patients. It will further gauge the potential role of somatic comorbidity and psychotropic polypharmacy, with emphasis on antipsychotics and antidepressants.

**Methods:** Density sampled case-control study of schizophrenic and bipolar patients from the Capital Region and Region Zealand in Denmark using prospective data from Danish registries and electronic medical records. Cases deceased between 1 January 2011 and 1 July 2016; controls were found by risk-set sampling, matched by date of birth +/- 180 days. We will use a fully Bayesian, non-linear, conditional logistic regression model to estimate the causal effect of serum sodium level on death, with covariates selected by a causal diagram. Continuous covariates will be modelled with restricted (natural) cubic splines to elicit non-linear relationships. A sample of 10 cases and 50 controls was drawn to ascertain that the proposed study be sound and realistic.

**Conclusion:** This study will expand the body of evidence on the effect of hyponatriaemia on mortality in schizophrenic and bipolar patients and may guide clinical decision-making in terms of both monitoring intensity and rational use of psychotropics.

## Introduction

In recent years several studies have shown that serum sodium in hospitalised somatic patients exhibits a j-shaped association with mortality, with lowest mortality in the normal range (138-142 mmol/L) [1-3]. Hypernatriaemia (high serum sodium) is rare and therefore a minor problem although associated with elevated mortality risk. In contrast, hyponatraemia is the most common electrolyte disorder in hospitalised patients; in a study of 53,236 persons from 2010 conducted in Boston, hyponatriaemia at admission (community-acquired hyponatraemia) occurred in 38% of hospitalisations and was associated with an adjusted odds ratio (OR) of 1.52 for in-hospital mortality [1]. Hospital-acquired hyponatremia developed in 38% of hospitalisations and was associated with an adjusted OR of 1.66 for in-hospital mortality [1]. The strengths of these associations tended to increase with hyponatraemia severity [1]. Another large cohort study from Boston produced similar estimates [2], and a meta-analysis of 15 studies indicated that any improvement of hyponatraemia were associated with a 43% reduced risk of overall mortality [3].

Patients with psychiatric diseases often develop hyponatriaemia, which could be due to a somatic comorbidity or to the syndrome of inappropriate secretion of anti-diuretic hormone (SIADH) induced by psychotropics. Psychiatric patients are more prone to somatic comorbidity than the general population, so in light of results from somatic patients, a key question is whether hyponatraemia in most patients is simply a marker of the severity of the underlying condition(s) that led to death or a direct contributor to death. Thus, this study will explore if the association between hyponatriaemi and mortality found in studies of somatic patients also exists in psychotic and bipolar patients, and in this context to understand the role of somatic comorbidity and psychotropic polypharmacy, with emphasis on antipsychotics and antidepressants.

## Methods

### Study design, data and patients

Register-based case-control study with patients from two Danish regions and prospective data from the National Patient Registry (NPR) with diagnoses and procedures from Danish (mainly public) hospitals; the national Cause of Death Register (CoDR) with death-related information; the Danish Civil Registration System (CRS) with demographic information; and in-hospital electronic medical records (EMRs) with drug administrations and biochemical analyses. NPR uses i.a. Danish versions of the 10th revision of the International Classification of Disease (ICD-10) and the Nordic Classification of Surgical Procedures (NOMESCO); drug administrations the Anatomic Therapeutic Classification (ATC); and biochemical values i.a. the Nomenclature for Properties and Units (NPU). CoDR and CRS use idiosyncratic encodings.

Drug administrations and biochemical data are timestamped with minute resolution; in-hospital drug administrations akin to ours have been validated [4], and biochemical samples are automatically timestamped at bed-side patient identification. NPR entries are timestamped with day resolution using the day of discharge, sufficient for identifying date of death and devise medical history covariates. Our biochemical data originate from two systems, BCC and Labka II, rolled out in our catchment area in 2010/11 and 2009/10, respectively. The registries go back decades [5]. The source population will consist of Danish residents with at least one ICD-10 F2 or F31 diagnosis from a hospital in Region Zealand or the Capital Region. Cases died in our catchment area between 1 January 2011 and 1 July 2016; 5 controls per case were found by risk-set sampling with replacement, matched on date of birth +/- 180 days [6]. We used the data from 10 risk sets (N = 60), picked at random, to ascertain that the planned analyses be appropriate and realistic, see table 1.

**Table 1.** Descriptive and summary statistics of 10 risk sets (60 patients). All variables, except age and sex, use a 5-year look-back window unless otherwise specified. Values are median (inter-quartile range) for numeric and N (%) for categorical variables.

| **Variable** | **Cases**<br>**(N = 10)** | **Controls**<br>**(N = 50)** |
| --- | --- | --- |
| Male sex | 8 (80%) | 16 (32%) |
| Age on index date | 71 (60–79) | 76 (65–86) |
| Mean p-sodium in mmol/L | 139 (132–143) | 140 (137–142) |
| Lowest p-sodium in mmol/L | 132 (129–140) | 136 (128–139) |
| Proportion of p-sodium measurements < 135 mmol/L | 20% (10%–90%) | 10% (0%–50%) |
| No. of p-sodium values per patient | 14 (2–63) | 15 (4–41) |
| Psychotropics used |  |  |
| Highest equivalent haloperidol dose | 5.0 (3.4–10.0) | 2.7 (0.9–19.6) |
| No. of antipsychotics (excl. lithium) | 2.0 (1–4) | 2 (1–5) |
| Highest equivalent fluoxetine dose | 23.5 (22.5–35.6) | 40.2 (22.2–44.4) |
| No. of SIADH-related antidepressants | 1 (1–2) | 1 (1–2) |
| Highest lithium dose | 0 (0–0) | 18.0 (9.0–18.0) |
| Years from first F2 diagnosis to index date | 13.9 (3.3–15.4) | 4.4 (1.9–16.7) |
| No. of distinct ICD-10 diagnoses recorded ever<sup>§</sup> | 28 (11–52) | 39 (15–65) |

<sup>§</sup> Denmark started using ICD-10 codes in 1994.

We refer the reader to the supplement which elaborates on and operationalises several methodological aspects of this study.

### Exposure and covariates

We use plasma sodium (p-sodium) status as exposure of main interest and operationalise this by the mean and minimum p-sodium concentrations (mmol/L) measured for each patient in the last five years before the index date. Because we expect a somewhat j-shaped relationship between these exposures and mortality risk and find parabolic functions overly rigid and unrealistic [7], we will use natural cubic regression splines to elicit non-linear relationships [6-9]. Natural cubic splines are flexible, smooth at the knots and fairly well-behaved in the tails, and they add relatively few extra parameters to estimate [10]. We will run a derived, secondary model in which splines are replaced by clinically meaningful bins compatible with the data—i.e. analysing the numeric variables as categorical—to facilitate clinical application and interpretation.

We have used DAGitty to devise the causal structure as a directed acyclic graph (DAG) that underpins our statistical model [11]. DAGitty evaluates if a causal effect of some exposure on an outcome be estimable with the available data, and identifies minimal sufficient covariate sets to estimate this effect, accounting for confounding without introducing biases. Our causal DAG (figure S1) yields one minimal sufficient covariate set (data types in parentheses): age (continuous), sex (female/male), highest dose of antipsychotics (continuous), number of antipsychotics (continuous), highest dose of SIADH-related antidepressants (continuous), number of SIADH-related antidepressants (continuous), highest lithium dose (continuous), schizophrenia diagnosis (nominal), bipolar diagnosis (nominal), presence of both schizophrenia and bipolar diagnosis (no/yes), CNS disease (no/yes), cardiovascular disease (no/yes), pulmonary disease (no/yes), malignant disease (no/yes), primary adrenal insufficiency (no/yes), renal disease (no/yes), surgery (no/yes). We use 5-year look-back window for all covariates.

### Data analysis

We will employ a conditional logistic regression model and use the R statistical programming language (www.r-project.org) for all analyses and visualisations. To, mainly, facilitate model validation and obtain intuitive uncertainty estimates, we will do full Bayesian parameter estimation using the *rstanarm* package [12]. Knots will be specified using quantiles [10] because their numbers are important but their exact locations less so [13]. To render estimation robust to outliers, the regression coefficients will have Student-t priors.

### Model diagnostics and performance

Parameter estimation with sampling requires some checks to ascertain the estimates’ credibility. We will run 4 Hamiltonian Monte Carlo chains with sufficient warm-up iterations and at least 1,500 effective posterior samples per parameter. Chain convergence will be checked visually (overlain trace and density plots) and numerically (potential scale reduction factor <1.05 for all parameters).

We will use posterior predictive checking to graphically verify that the model adequately describe the input data, and Pareto-smoothed importance sampling leave-one-out cross-validation (PSIS-LOOCV) to check its out-of-sample predictive performance and model comparison [14]. PSIS-LOOCV is an approximation method, and we will do genuine K-fold cross-validation should it fail.

### Sensitivity analyses

We will test our findings’ robustness to select design decisions and discuss relevant implications: mean p-sodium weighted by temporal contribution instead of crude means, medical history as count variables instead of binary, psychotropic prescriptions instead of administrations, and Gaussian priors instead of Student t-distributed ones. Further, to assess the potential effect of truncation bias introduced by using 5-year mean and minimum p-sodium, we will also run the model with a 1-year look-back window.

### Reporting

The results of this study are to be published in an international peer-reviewed journal, observing the STandardising Reporting of OBservational studies in Epidemiology (STROBE) and REporting of studies COnducted using Routinely-collected health Data (RECORD) guidelines, and disclose all code used to carry out the analysis. To give due emphasis to the main results of the study, the report will feature effect-size estimates of p-sodium status, antidepressants and antipsychotics graphically because numerical summaries of splines are difficult to intuit. We will keep the covariate estimates out of the main text to avoid the table 2 fallacy [15] but produce a supplement containing i.a. medians and 95% credible intervals of all regression coefficients and model diagnostics. The posterior samples will be made publicly available for scrutiny and potential repurposing in future Bayesian studies.

### Ethics

The data live in a secure cloud on a high-performance computing infrastructure (Computerome) in Denmark. Data collection and linkage were approved by the Danish Health Data Authority (FSEID-00003724) and the Danish Patient Safety Authority (3-3013-1731-1). No additional data were or will be collected, obviating ethics committee approval. The study has been registered at ClinicalTrials.gov (NCT04409626).

## Discussion

With this study, we aim to assess if hyponatraemia in schizophrenic and bipolar patients increases the risk of dying. The relationship between hyponatraemia and mortality is widely studied in somatic populations, but the body of knowledge on psychiatric patients is limited [16]. SIADH-induced hyponatraemia, however, is a well-known adverse effect of antipsychotic treatment, so we chose to study schizophrenic and bipolar patients because long-term or even lifelong antipsychotic treatment is common among these patients. To avoid prespecifying a hyponatraemia threshold, we will model the mean and minimum p-sodium with natural cubic splines to elicit non-linear relationships.

This study will feature some important strengths. First, we combine registry and clinical data to yield a comprehensive picture of the included patients. Many large-scale pharmacoepidemiological studies use registries only, precluding studying hypotheses as the one we advance here. Second, the statistical model is built on a causal structure, ensuring that—conditional on that structure—the estimated effect of p-sodium status on mortality risk is causal rather than associational. Large-scale, unsupervised propensity score methods have gained much traction in recent years but are arguably inferior to explicit causal modelling, because they cannot guarantee that estimated effects be causal. Indeed, identifying a sufficient set of covariates requires domain knowledge and cannot be done by statistical methods alone [17]. Third, we employ Bayesian parameter estimation to obtain intuitive and probabilistic results: posterior distributions reflect our uncertainty about the estimates of interest. This property is often and erroneously ascribed to traditional confidence intervals although they actually reflect the intangible notion of infinite series of repeated experiments. Also, Bayesian estimation facilitates the creation of complex models [18], explicates distributional assumptions, and allows for building pre-existing knowledge into the model. Fourth, we elicit non-linear effects of p-sodium and polypharmacy covariates with natural cubic splines. Finally, several sensitivity analyses will elucidate if and how study design aspects affect the parameter estimates.

Notwithstanding these strengths, the study will have some limitations. First, our causal structure may be incomplete, which could bias our causal effect estimates. Disagreement on the causal structure, and the consequent impact on the findings, is amenable to further scrutiny by comparing the results of running the analyses anew with alternative covariate structures and formal tests provided by DAGitty [11]. Second, there could be unobserved periods of hyponatraemia, and systematic differences between groups would introduce bias. However, p-sodium is a cheap routine analysis, so we do not expect such misclassification of exposure to be important. Third, we use non-informative priors, but these should be dominated by the data and inference on their effect sizes is not of interest because they serve as nuisance parameters [19]. Fourth, we cannot link our in-hospital records to medicine dispensations at community pharmacies, but psychotic and bipolar patients tend to be followed closely at the hospital, and because doctors are obliged to update patients’ medicine profiles in the EMR during such visits, we anticipate realistic data on psychotropics use and will assess this with a sensitivity analysis. Finally, equivalent-dose conversion of antidepressants and antipsychotics will surely be imprecise, but converted doses are reasonable in the sample.

We believe this study will expand the body of evidence on the effect of hyponatraemia on mortality in schizophrenic and bipolar patients and may guide clinical decision-making in terms of both monitoring intensity and rational use of psychotropics.

## Data Availability

We use sensitive data from national registries and electronic medical records. We cannot share these.

## Funding

Innovation Fund Denmark (5153-00002B). Novo Nordisk Foundation (NNF14CC0001).

## Study registration

ClinicalTrials.gov ID: NCT04409626

## Contributions (cf. the <u>CRediT</u> contributor roles taxonomy)

Conceptualisation: BS, NG, GJ. Data curation: BS, SB. Formal analysis: BS, NG, SA, LT, GJ. Methodology: BS, NG, SA, LT, GJ, Software: BS. Drafting: BS, SA, GJ. Funding acquisition: SA, SB. Resources: SB. Resources: SB. Supervision: SA, SB, GJ. Review: All.

## Conflicts of interest

The authors declare no conflicts of interest.

## Notes

### Competing Interest Statement

The authors have declared no competing interest.

### Clinical Trial

NCT04409626

### Funding Statement

The study was funded by Innovation Fund Denmark (5153-00002B) and Novo Nordisk Foundation (NNF14CC0001).

### Author Declarations

Data collection and linkage were approved by the Danish Health Data Authority (FSEID-00003724) and the Danish Patient Safety Authority (3-3013-1731-1).

